# Longitudinal assessment of functional antibodies to a novel influenza virus strain across age groups

**DOI:** 10.64898/2026.02.21.26346781

**Authors:** Anke Huckriede, Ilse Hoorn, Manas Joshi, Jacqueline de Vries-Idema, Gestur Vidarsson, Puck B. van Kasteren, Martin Beukema

**Author notes:** Corresponding author: Anke Huckriede.

## Abstract

Newly emerging influenza virus strains pose a constant threat as they encounter a population lacking neutralizing antibodies against the new strain. However, cross-reactive non-neutralizing antibodies (nnABs) may be present and assist in mitigating disease symptoms via various effector mechanisms, including antibody-dependent cellular cytotoxicity (ADCC). Although nnABs to influenza virus have received more attention lately, little information is available on their age-related prevalence, steady-state levels, functional properties, and changes in these parameters over time.

Using longitudinal samples from adolescents, adults, and older adults, collected before and after the 2009 swine flu pandemic, we comprehensively characterized the specificity and functionality of nnAB responses against H1N1 pandemic 2009 (H1N1pdm09) virus. Remarkably, all participants exhibited cross-reactive antibodies to this virus before having encountered it through infection or vaccination, with the highest baseline levels observed in older adults. The levels of these IgG antibodies showed a strong correlation with engagement of fragment crystallizable γ receptor IIIa (FcγRIIIa) and ADCC activity, both of which were notably lower in adolescents compared to adults and older adults. Without infection or vaccination, average amounts of H1N1pdm09-reactive antibodies remained relatively stable on population level over the 5-year study period. However, on an individual level, substantial increases and decreases occurred. H1N1pdm09 infection or vaccination significantly enhanced specific antibody levels and the FcγRIIIa-engaging capacity of these antibodies in all age groups. ADCC-mediating antibodies increased however only in adolescents, reaching the same level as observed in the adult groups.

Taken together, our results demonstrate the presence of cross-reactive, non-neutralizing, functional, and boostable antibodies against a never-encountered influenza virus strain across all age groups. These antibodies can potentially contribute to protection from severe disease. Accordingly, in case of a newly emerging virus, their further enhancement by vaccination could be beneficial as an immediate protective measure before a strain-specific vaccine becomes available.

**Author summary:** Nearly everyone has contracted influenza and/or has been vaccinated against influenza several times over the years. While the antibodies raised during these earlier encounters will not prevent infection by a newly emerging influenza virus strain, they can help to protect from severe disease. Therefore, it is important to determine the prevalence and quantity of these antibodies, understand their mechanisms of action, assess their persistence over time, and examine potential age-related differences in these parameters. We studied antibody responses to the H1N1pdm09 virus in blood samples of young, adult, and older adult individuals from a large cohort study. Irrespective of age, all blood samples contained antibodies that reacted with a never-before-encountered influenza virus strain. The amounts of these antibodies were initially lower in adolescents but with time increased, reaching the same levels as observed in adults. Importantly, infection with or vaccination against the new virus strengthened the responses in all age groups. We conclude that boosting such broadly-reactive antibodies through vaccination could serve as an immediate strategy when a new virus emerges, buying critical time to develop a more specific vaccine.

## Introduction

The ongoing circulation of H5N1 influenza virus strains in poultry worldwide and in dairy herds in the USA, the recent reports on fatal human H5Nx infections from Cambodia and the USA, and the regular notifications about human infections with other influenza virus strains underline the constant threat of a zoonotic influenza virus strain crossing the species barrier and causing a next pandemic (1). Such an emerging influenza virus could spread rapidly since the population does not possess neutralizing antibodies against the new strain. However, cross-reactive non-neutralizing antibodies (nnABs) might exist and contribute to protection from severe disease by effector mechanisms involving the fragment crystallizable (Fc) domain of the antibody molecules (2–5).

Non-neutralizing antibodies (nnABs) have long been neglected but are currently gaining attention due to emerging evidence of their role in protecting against severe disease caused by SARS-CoV-2 and influenza viruses (6–9). Importantly, many nnABs to the influenza virus recognize conserved viral proteins or protein domains and are accordingly cross-reactive with a broad range of influenza virus strains (10). Effector mechanisms engaged by nnABs comprise antibody-dependent complement deposition (ADCD), antibody-dependent phagocytosis (ADCP), and antibody-dependent cellular cytotoxicity (2–4), with ADCC most closely associated with protection from symptomatic influenza virus infection (11). ADCC involves the binding of antibodies to cell surface-exposed viral proteins on infected cells, followed by recognition of the opsonized cells by the IgG-Fc receptor IIIa (FcγRIIIa) on natural killer (NK) cells and release of perforin and granzyme that kill the infected cell (12). Among the four human IgG subclasses, only IgG1 and IgG3 have a strong affinity for binding to FcγRIIIa, with IgG1 being by far the most abundant IgG subclass (13). FcγRIIIa-binding is further determined by the core glycosylation pattern of the antibodies: non-fucosylated IgG1 binds to the receptor with strongly enhanced affinity compared to fucosylated IgG1 (14). Although the majority of immune responses involves fucosylated IgG, varying amounts of specific afucosylated IgG have been observed in alloimmune responses, responses to erythrocyte antigens of malaria, but also to glycoproteins of several enveloped viruses (15,16).

Despite the recognized importance of nnABs for mitigation of influenza in humans, little information is available on the prevalence, durability, and functional performance of cross-reactive nnABs in the general population and how these parameters might be influenced by age. We earlier studied immunity to 5 different influenza virus strains in serum samples from young, middle-aged, and older individuals taken with a 5-year interval (17). For this purpose, we made use of the Lifelines biobank, a 3-generation longitudinal population-based cohort study running in Groningen, The Netherlands (18). The samples were collected before (adults, older adults) or around (adolescents) the 2009 swine flu pandemic and 5 years later. Our results revealed that neutralizing antibodies in each age cohort preferentially bound to a virus strain the cohort was exposed to early in life or for a long period of time. In contrast, virus-binding antibodies, measured by ELISA, showed no strain-specific preferences and were of similar magnitude in all age groups. Interestingly, such antibodies cross-reacted with virus strains the participants had never been exposed to, including the H1N1pdm09 strain A/California/07/2009 (A/Cal in the following) (17).

Here, we study the H1N1pdm09-reactive antibodies in more detail, determining antibody targets, antibody subclass profiles, FcγRIIIa-binding, and NK cell activation, and assessing the changes in these parameters over the 5-year sampling interval. Our results demonstrate the presence of antibodies reactive with H1N1pdm09 but not necessarily targeting the surface proteins hemagglutinin (HA) or neuraminidase (NA), before exposure to the virus or vaccine in all age groups. Importantly, older individuals displayed similar or even higher levels of H1N1pdm09-binding and ADCC-mediating antibodies as/than adults and adolescents. Young individuals initially had the lowest antibody titers but showed the most pronounced increases in virus-binding and ADCC-mediating antibodies over time.

## Results

### H1N1pdm09-reactive antibodies are present in pre-pandemic serum samples

In our previous study, we used serum samples from 60 adolescents, 60 adults, and 60 older adults (referred to as seniors in the following), collected at two assessment moments (A1 and A2), five years apart (17). For the current study, we selected from these serum samples those with a microneutralization (MN) titer ≤ 40 for H1N1pdm09 at assessment A1. This enabled us to measure cross-reactive antibody levels to H1N1pdm09 in presumably unexposed individuals. Focusing on sero-negative samples led to the exclusion of 16 out of 60 samples from adolescents and 5 out of 60 samples from middle-aged adults (referred to as adults in the following) (Fig. 1). Since the A1 samples from adolescents were taken in 2011/12, the MN-positive individuals in this age group had likely been infected by H1N1pdm09 virus or had been vaccinated. However, the A1 samples of the adults were taken early in 2009, making exposure to an H1N1pdm09 virus very unlikely. The origin of MN antibodies in these samples is thus unclear. No MN-positive sera were identified in the samples from seniors.

**Fig. 1:**
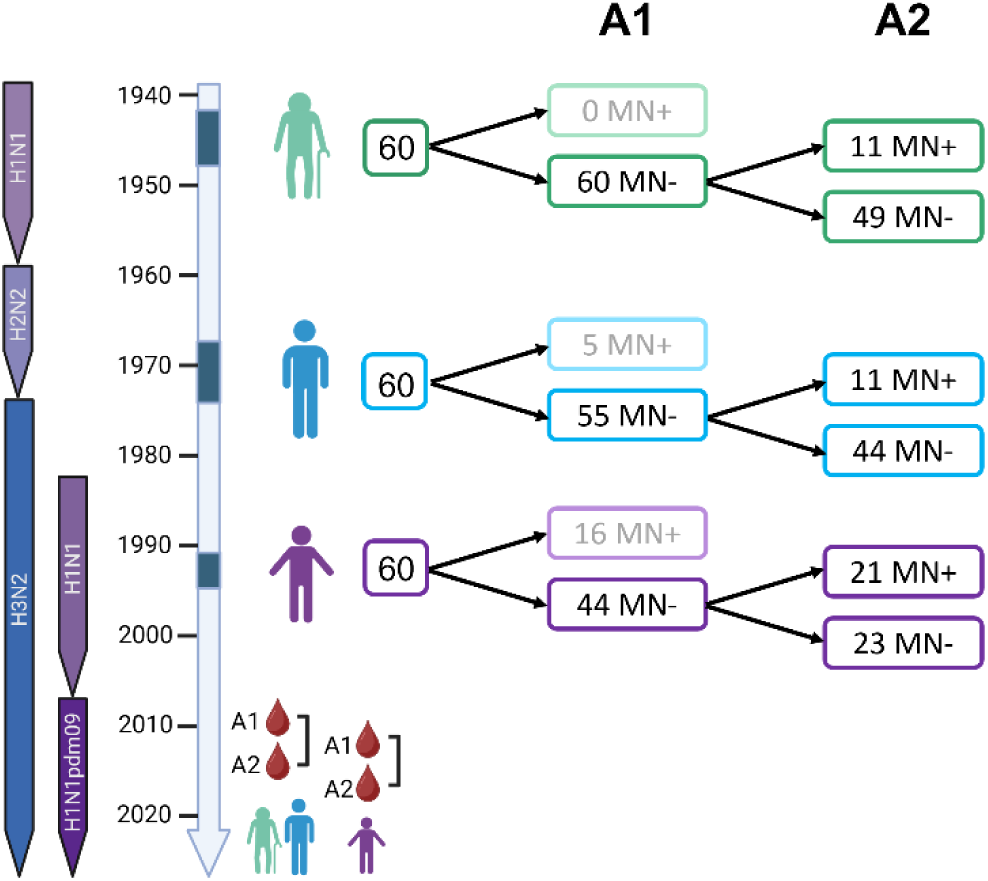
Overview of serum samples used in the study. Serum samples from 60 seniors, 60 adults, and 60 adolescents were collected in 2009 and 2014 from adults and seniors and in 2011-12 and 2015-17 from adolescents (birth periods indicated by dark blue boxes, sampling moments indicated by diamonds and blood droplets on the timeline). The samples were classified as MN+ or MN- if their microneutralization antibody titer to H1N1pdm09 was ≥80 or ≤40, respectively (determined in Sicca et al 2022). Only A1 MN- samples were included in the current study and were again classified as MN+ and MN- based on their MN titer at A2.

To reconfirm the presence of H1N1pdm09-binding antibodies and to identify the targets of these antibodies, we performed ELISAs on the serum samples collected at A1, coating the plates with whole inactivated virus (WIV), HA subunit vaccine (SU), or nucleoprotein (NP), or used data from an enzyme-linked lectin assay (ELLA) to measure neuraminidase (NA)-inhibiting antibodies performed earlier (17) (Fig. 2). Our results demonstrate the presence of antibodies reactive with H1N1pdm09 WIV, before exposure to the virus or vaccine, in all participants, with the highest levels observed in seniors (Fig. 2A). All participants also possessed NP-binding antibodies which were of similar magnitude in all age groups (Fig. 2D). In contrast, antibodies to SU and NA were undetectable in some participants, and levels of NA-inhibiting antibodies were significantly lower in adults than in adolescents and seniors (Fig. 2B, C). Taken together, these results reveal a high prevalence of antibodies cross-reactive with diverse proteins of a never-encountered virus strain in all studied age groups.

**Fig. 2:**
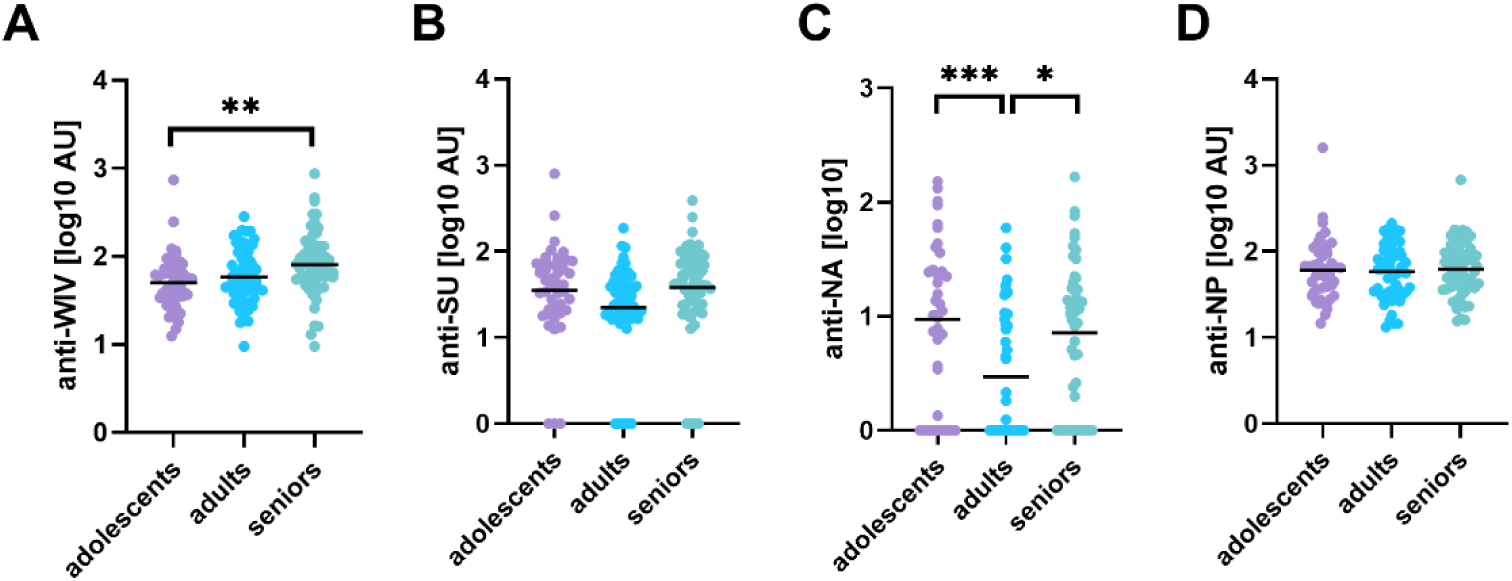
Influenza-reactive antibodies vary with age. Serum samples of adolescent (n=44), adult (n=55), and senior (n=60) individuals, negative for neutralizing antibodies to H1N1pdm09, were used to determine antibodies to H1N1pdm09 whole inactivated virus (A), subunit (B), or NP (D) by ELISA, and antibodies to NA by ELLA (C). Antibody levels measured by ELISA are depicted as arbitrary units (AU) calculated using a calibration set of sera. If the OD value of a prediluted serum sample was lower than the OD value of the highest dilution of the calibrator the sample was assigned an AU value of 0. Levels of NA-inhibiting antibodies are depicted as log10 of the serum dilution resulting in 50% inhibition of NA activity. Average values are indicated by a horizontal line. Statistical differences were calculated by ordinary one-way ANOVA with Tukey’s multiple comparison test. * p<0.05, ** p<0.01, *** p<0.005.

### Functional antibody responses to H1N1pdm09 are maintained across age groups

Next, we determined the antibody subclass profiles of H1N1pdm09-reactive antibodies, as these can be of relevance for the functional performance of nnABs. Overall, we found little differences in the levels of WIV-binding IgG1, IgG2, and IgG4 among the age groups, except for a trend to slightly lower IgG1 and IgG2 in adolescents and lower IgG4 levels in seniors (Fig. 3). IgG3 could not be detected in any serum samples at the used serum predilution of 1:50 (results not shown).

**Fig. 3:**
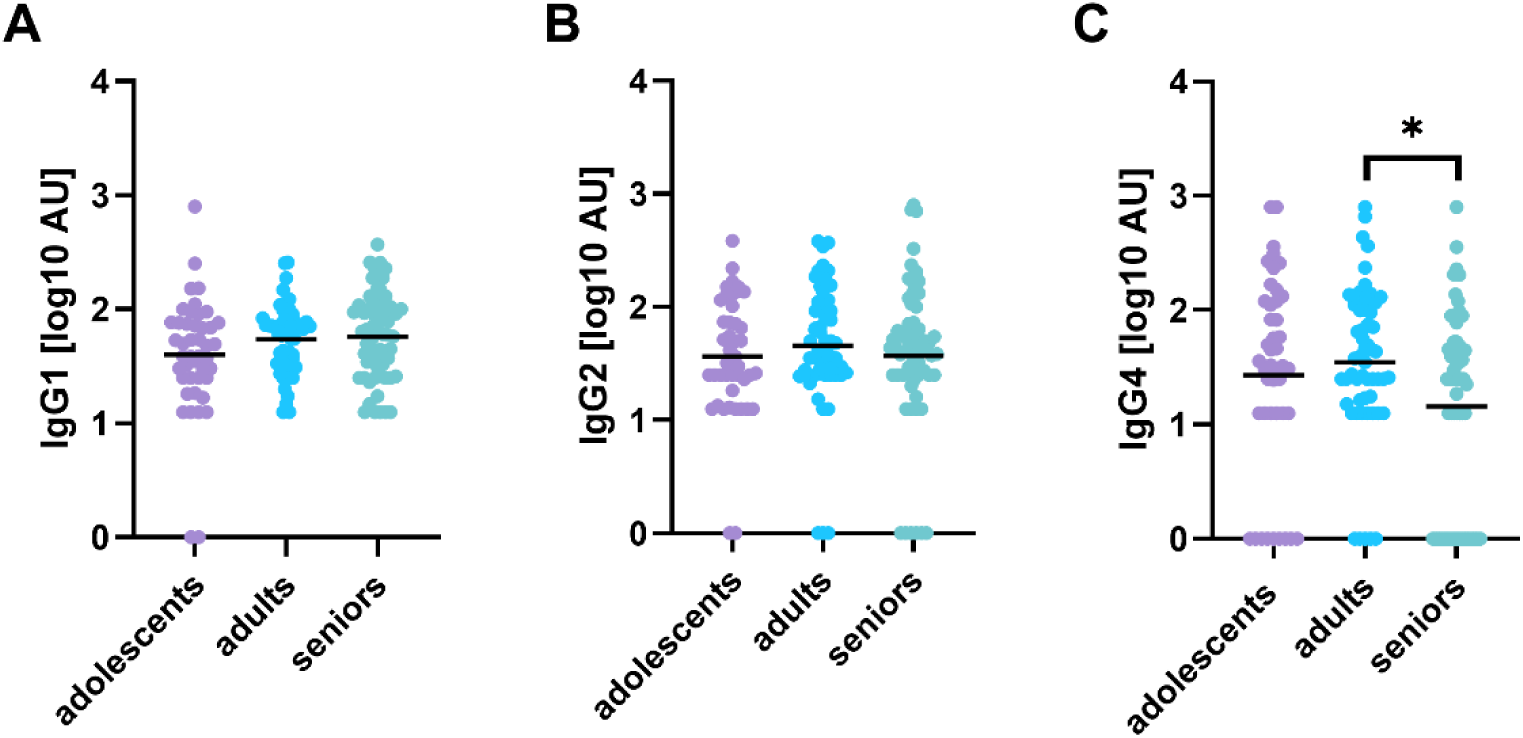
Minor variation in IgG subclass distribution of H1N1pdm09-specific antibodies with age. Using ELISA plates coated with H1N1pdm09 WIV, IgG1 (A), IgG2 (B), and IgG4 (C) levels were quantified in the sera of adolescent, adult, and senior individuals. All included participants were seronegative for neutralizing antibodies to H1N1pdm09. Antibody levels are depicted as arbitrary units (AU) calculated for each subclass using a calibration set of sera. If the OD value of a prediluted serum sample was lower than the OD value of the highest dilution of the calibrator the sample was assigned an AU value of 0. Average values are indicated by a horizontal line. Statistical differences were calculated by ordinary one-way ANOVA with Tukey’s multiple comparison test. * p<0.05.

To get insight into the functional performance of the antibodies to H1N1pdm09, we measured the levels of FcγRIIIa-engaging antibodies, which are sensitive to both quantity and fucosylation status of IgG1, and determined the potential of the antibodies to activate NK cells to express CD107a as a proxy for ADCC (Fig. 4) (12,14, 15). Despite the absence of age-related differences in the levels of IgG1, we observed that levels of H1N1pdm09-reactive FcγRIIIa-engaging antibodies were lower in adolescents than in adults (significant) and seniors (non-significant), the latter two groups exhibiting about equal levels of these antibodies. In contrast, no age-related differences were found in the percentages of CD107a-expressing NK cells after exposure to antibody-opsonized WIV, indicating that antibodies from the different age groups had equal potential to trigger ADCC.

**Fig. 4:**
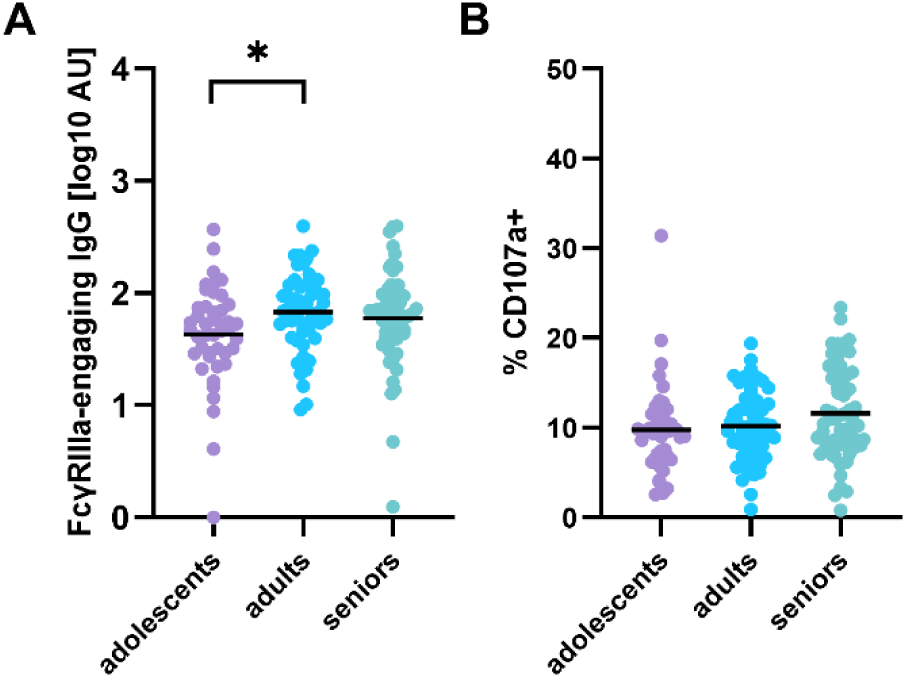
Age moderately affects antibody effector functions. The sera of adolescents, adults, and seniors were tested for H1N1pdm09-reactive antibodies engaging FcγRIIIa (A) and antibodies activating NK cells (B). Results are expressed as arbitrary units (AU) for FcγRIII-engaging antibodies and as percentage of CD107a-positive NK cells for NK cell-activating antibodies. Statistical differences were calculated by ordinary one-way ANOVA with Tukey’s multiple comparison test. * p<0.05.

Correlation analyses over the entire study population revealed significant correlations between total IgG and all 3 measured IgG subclasses (Fig. 5A). Furthermore, FcγRIIIa-binding and NK cell activation correlated strongly with each other and with total IgG and IgG1 but not with IgG2 and IgG4 (Fig. 5A). Investigating these correlations per age group revealed that adolescents showed particularly strong correlations of total IgG with IgG subclasses and of IgG and IgG1 with FcγRIIIa-binding and NK cell activation (Fig. 5B). In adults and seniors, these correlations were weaker or absent (Fig. 5C, D). This was due to some individuals with rather moderate IgG and IgG1 titers but high levels of FcγRIIIa-engaging antibodies; such individuals were not found in the adolescent group.

**Fig. 5:**
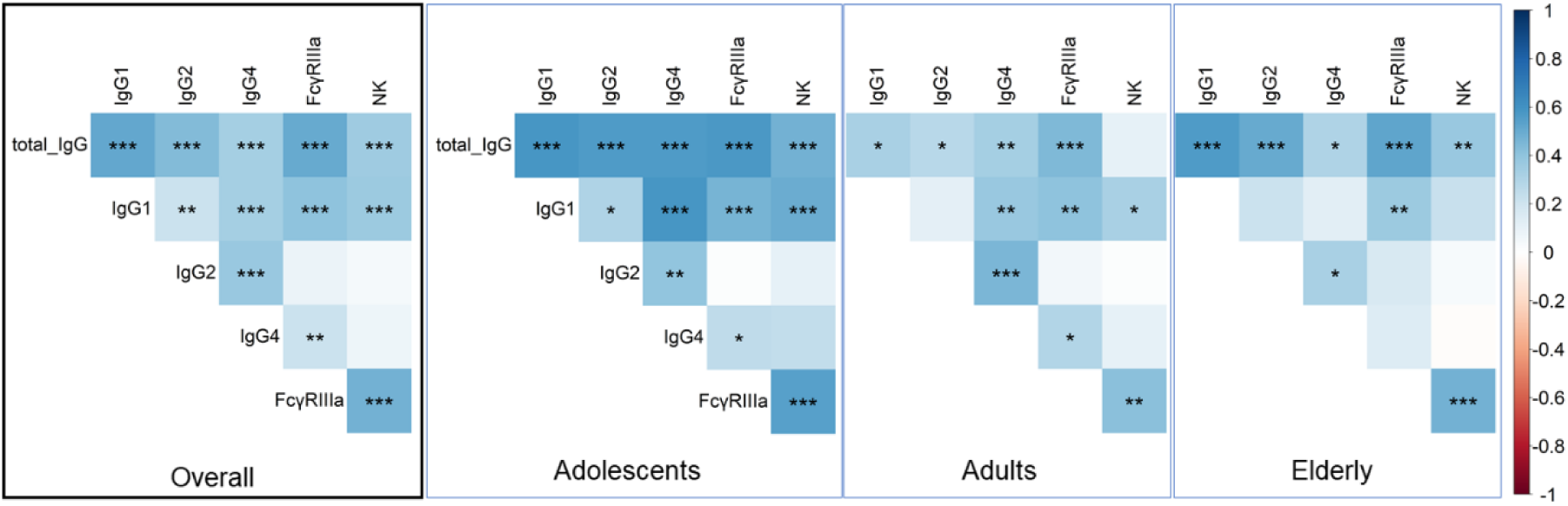
IgG and IgG1 correlate with each other and with nnAB effector functions across all age groups. Correlation analysis of total IgG, IgG subclasses, and functional antibody properties for all participants (A) and separate for adolescents (B), adults (C), and seniors (D). The colour indicates the level of correlation and the stars indicate the level of significance with * p<0.05, ** p<0.01, and *** p<0.001.

### Longitudinal analysis reveals age-dependent dynamics of antibody responses to H1N1pdm09

The longitudinal setup of the Lifelines cohort study enabled us to investigate the dynamics of the different antibody responses on a population and individual level over time. In adolescents, antibodies to WIV, SU, and NA increased significantly between A1 and A2, while antibodies to NP did not change. In contrast, adults and seniors only displayed significant changes in antibodies to SU but not to the other antigens (supp. Fig. 1). Increases in antibody levels were mainly driven by exposure to virus or vaccine as became evident when we analyzed data of individuals who did (MN+) and did not (MN-) seroconvert between A1 and A2 (Fig. 6). MN+ individuals showed significant increases of antibodies to WIV, SU, and NA but not NP (Fig. 6) and > 2-fold increases were observed in most of these participants (supp. Fig. 2). In contrast, in MN- individuals, significant changes were found only for SU-reactive antibodies and only in adults and seniors (Fig. 6). Nevertheless, more than 2-fold increases and decreases in antibody levels were observed in 25-70% of the MN- individuals, especially for SU- and NA-reactive antibodies (supp. Fig. 2) indicating substantial dynamics of the antibody repertoire.

**Fig. 6:**
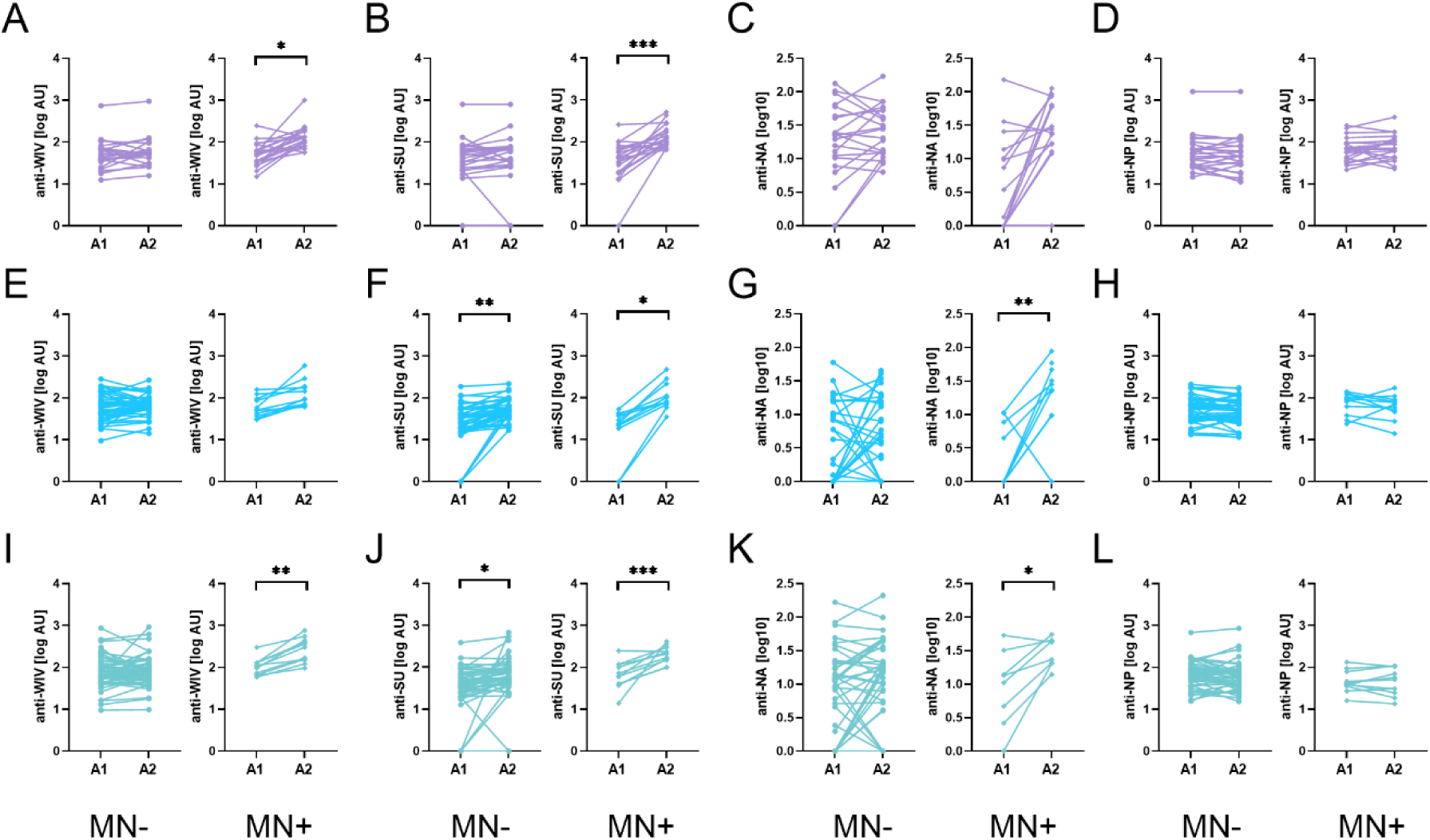
Increases in antibody titers to WIV, SU, and NA are driven by individuals who were exposed to the virus or vaccine between A1 and A2. Changes in antibody levels are displayed for individuals not exposed (MN-) or exposed (MN+) to H1N1pdm09 virus or vaccine between A1 and A2 for adolescents (A-D), adults (E-H), and seniors (I-L). Significant increases or decreases in antibody levels from A1 to A2 determined by Welch’s T test are indicated by * p<0.05, ** p<0.01, and *** p<0.005.

In line with increases in H1N1pdm09-reactive antibodies, FcγRIIIa-engaging antibodies increased in adolescents but not in adults and seniors between A1 and A2 (supp. Fig. 3 left). When analyzed separately for MN- and MN+ individuals, significant increases were observed only in MN+ individuals (Fig. 7) with > 55% of these individuals displaying >2-fold increases (supp. Fig. 4A). Yet, frequent changes in the levels of FcγRIIIa-engaging antibodies were also found in non-exposed (MN-) individuals (supp. Fig. 4A). The increases in FcγRIIIa-engaging antibodies did not affect the levels of NK cell-activating antibodies which were similar across the age groups at both assessments (supp Fig. 3 right). Only in MN+ adolescents, NK cell-activating antibodies increased significantly (Fig. 7). Nevertheless, the levels of these antibodies changed by more than 2-fold in at least 50% of MN+ and MN- individuals of all age groups indicating common fluctuations in antibody steady state levels (supp Fig. 4B).

**Fig. 7:**
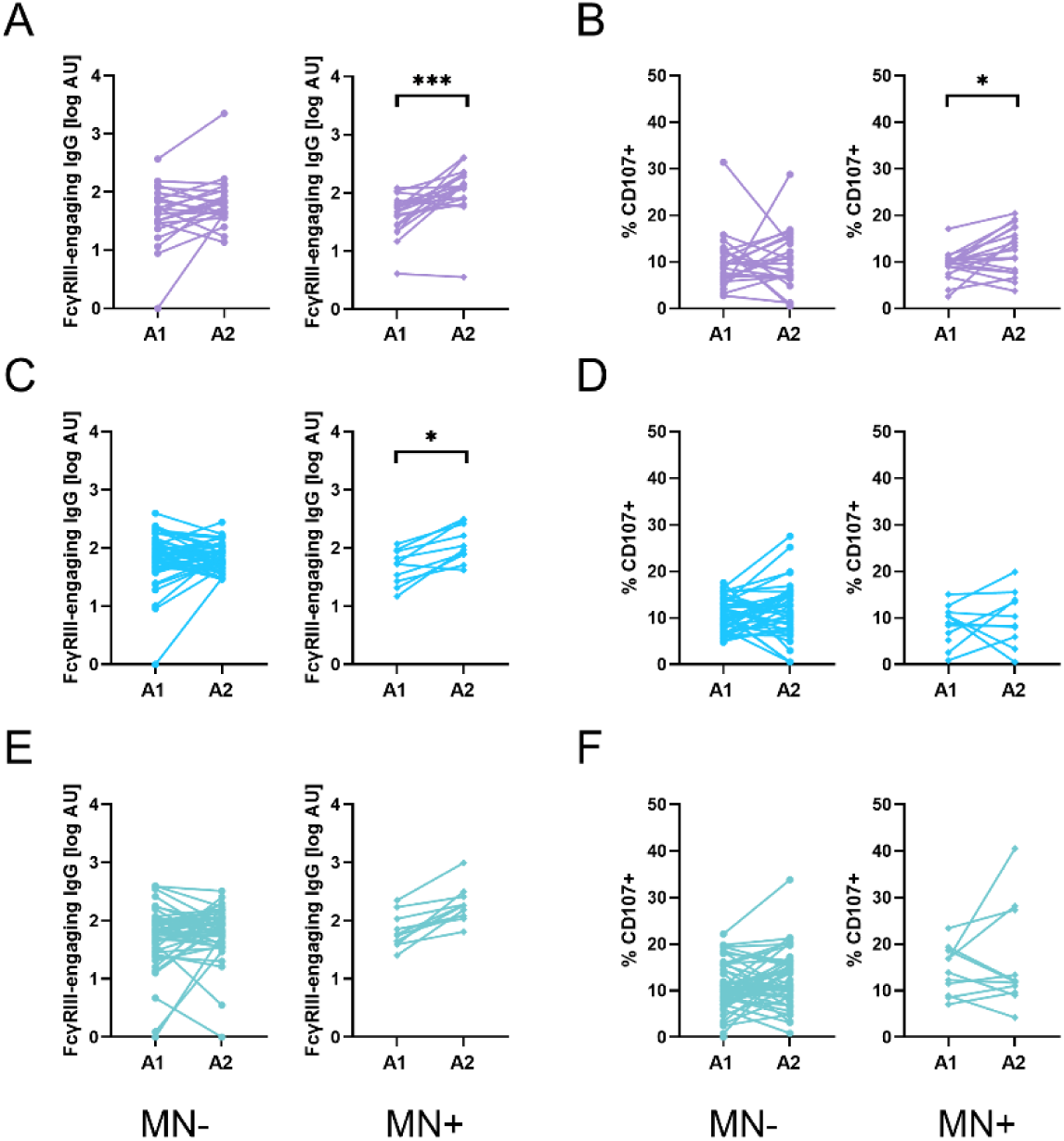
Changes of FcγRIIIa-engaging antibodies but not NK cell-activating antibodies are pronounced in H1N1pdm09-exposed individuals. FcγRIIIa-engaging antibodies (A, C, E) and NK cell-activating antibodies (B, D, F) were measured for samples taken at A1 and A2 from H1N1pdm09 non-exposed (MN-) and exposed (MN+) individuals. Changes in antibody levels over time are depicted for adolescents (A, B), adults (C, D), and seniors (E, F). Significance of differences in titers between A1 and A2 were assessed by one-way ANOVA with Šídák’s multiple comparison test. * p<0.05, *** p<0.005.

Taken together, these results demonstrate that without exposure to the virus or vaccine, average antibody levels were stable on a population level but not on an individual level where substantial increases and decreases were observed. Exposure to H1N1pdm09 virus or vaccine boosted antigen-binding and functional antibodies in all age groups but most strongly in young individuals.

## Discussion

In this study, we investigated nnABs reactive with antigens of A/Cal, an H1N1pdm09 virus strain, in young, adult, and senior cohorts. Even before exposure to H1N1pdm09 antigen through infection or vaccination, all participants had antibodies reactive with the virus, with the highest titers observed in seniors. IgG titers correlated with levels of FcγRIIIa-engaging and NK cell-activating antibodies, which were equally high or higher in seniors and adults than in adolescents. Without exposure to H1N1pdm09 virus or vaccine, antibody titers in the studied age groups were rather stable over the 5-year study period, although on an individual basis substantial increases and decreases occurred. Exposure significantly enhanced antibody levels to WIV, SU, and NA, but not NP, and boosted FcγRIIIa-binding antibodies, especially in adolescents.

Our results reveal a high prevalence of antibodies reactive to a virus the participants had presumably never encountered, as demonstrated by the absence of neutralizing antibodies. As such, they confirm earlier observations on H1N1pdm09-reactive antibodies measured by ELISA in individuals seronegative for hemagglutination-inhibiting (HI) antibodies to this virus (19–21). Most probably, the H1N1pdm09-binding antibodies were induced by earlier infection or vaccination and are directed to conserved proteins or protein domains. Indeed, conserved domains, located in the HA stem and – to a lesser extent - the HA head region, NA, M1, M2, and NP, have been identified as targets of nnAbs to influenza virus (22,23). In line with that, we found antibodies cross-reactive with WIV or the highly conserved NP in all participants. In contrast, antibodies to the more variable surface proteins HA and NA were detected in most but not in all individuals. This might reflect more frequent boosting of antibodies to the most conserved viral proteins.

With few exceptions, all sera contained H1N1pdm09-reactive FcγRIIIa-engaging and NK cell-activating antibodies. Many studies have reported on ADCC-mediating antibodies to H1N1pdm09 virus in sera devoid of HI or MN antibodies (24–27). It has long been speculated that these antibodies contribute to protection from severe disease (24); evidence for this hypothesis is recently accumulating (24,25,27). In a controlled human infection model of influenza, high ADCC antibody titers were found to be associated with reduced disease symptoms (28). Moreover, a recently published post-hoc study identified antibody-mediated NK cell activation, being the mechanism behind ADCC, as an independent correlate of immunity against influenza infection (11). These results highlight the relevance of immune mechanisms beyond antibody-mediated neutralization for the control of influenza, and our study confirms that such mechanisms are present in a large part of the population across all age groups.

Exploiting samples taken about 5 years after the first blood draw, we were able to follow antibody steady-state amounts in the same individuals over time. On a population level, we observed stable or increasing levels of antibodies to the different virus proteins, paralleled by stable or slightly increasing levels of FcγRIIIa-binding and NK cell-activating antibodies. While the durability of neutralizing antibodies has been investigated frequently, few studies have assessed influenza-specific IgG levels over time. Miller and co-workers measured IgG responses to various influenza virus strains in longitudinal samples taken at 5-year intervals over a 20-year period (29). In line with our results, they observed stable or slightly increasing IgG titers, depending on the virus strain studied. Similarly, Jia et al reported on stable titers of H1-binding IgG over a 5-year period in children (25). In the same samples used for the current study, we previously found that the antibody titers to three different H1N1 and H3N2 influenza virus strains from 1934, 1968, and 1999 remained stable or increased between the two assessments (17). Only IgG reactive with A/Perth/16/2009 (H3N2), the strain most recently encountered by the study participants, declined slightly. Previous studies on the maintenance of ADCC-mediating antibodies revealed stable titers over time, irrespective of exposure to viral antigen (25). This is in line with our results for adults and seniors, while in exposed young individuals, we observed significant increases in FcγRIIIa-engaging and NK cell-activating antibodies. Most likely, regular boosting by infection or vaccination during childhood and adolescence results in stable levels of functional IgG once adulthood is reached.

Despite relatively stable antibody levels on the population level, we observed substantial increases and decreases of H1N1pdm09-reactive antibodies on an individual level. Net increases in antibody titers were seen mainly for antibodies to the surface proteins HA and NA and were driven by individuals exposed to H1N1pdm09 virus or vaccine between the two sampling moments. However, increases in antibodies to HA were also observed in some adults and seniors who did not show significant increases in neutralizing antibodies. From the available data, it cannot be deduced whether these participants were infected or vaccinated with H1N1pdm09 but did not seroconvert, or whether the HA-reactive antibodies were enhanced by exposure to another virus strain. Nevertheless, this observation indicates that cross-sectional samples can give information on the prevalence of partially protective antibodies in the population but should not be used to predict protection on an individual level.

Importantly, we did not observe overt differences in the prevalence or the levels of H1N1pdm09-reactive antibodies in pre-pandemic samples from the different age groups, except for somewhat higher WIV-reactive antibody levels in seniors and lower levels of HA- and NA-reactive antibodies in adults. The latter observation could be the result of less frequent infections or vaccinations of adults compared to adolescents and seniors, respectively. Overall, compared to adolescents, adults and seniors showed more variation in responses and a weaker correlation between IgG and IgG subclasses on the one hand, and IgG, FcγRIIIa-binding, and NK cell-activating antibodies on the other hand. This points to a general broadening of the responses with increasing age due to accumulating exposures to (different) influenza antigens.

Interestingly, the levels of ADCC-mediating antibodies were higher in pre-pandemic samples of adults and seniors than of adolescents, although significance was reached only for FcγRIIIa-engaging antibodies. Other authors also reported on higher prevalence and/or levels of ADCC-mediating antibodies with advancing age (19,24). Moreover, the antibody responses to H1N1pdm09 were found to be broader and of higher affinity in old than in young individuals (30), pointing to maturation of the immune response to influenza virus with more frequent exposures. Between the two assessments, target protein-binding and functional antibodies changed most strongly in adolescents, bringing the levels of these antibodies to equal heights as observed in adults and seniors. These latter two groups showed little changes in binding and functional antibodies, except for increasing antibodies to HA. However, when exposed to H1N1pdm09 virus or vaccine, significant increases in virus-binding and ADCC-mediating antibodies were observed in adults and seniors, testifying successful boosting of memory responses.

The strength of our study is the assessment of multiple antibody response parameters in longitudinal samples from well-defined age groups of substantial size. Very few longitudinal studies on non-neutralizing influenza-specific antibodies have been published, and those that have are often not age-stratified or involve only a few study subjects (25,29). Here, our study fills an important knowledge gap by providing comprehensive data. Limitations of the study are the rather long interval of 5 years between the two blood draws and the lack of information on infections or vaccinations in between. To overcome this shortcoming in the future, participants of long-term cohort studies, as Lifelines, should be asked to provide this information annually via questionnaires, since more frequent sampling will often be unfeasible.

Taken together, our results confirm the high prevalence of functional non-neutralizing antibodies cross-reactive with a never-encountered virus strain described earlier. Furthermore, they demonstrate that the levels and dynamics of these antibodies, before and after antigen encounter, are rather similar across different age groups, with changes over time being most distinct in young individuals. Importantly, ADCC-mediating antibodies do not decline with time or increasing age and can contribute to disease mitigation in case of infection. Given the high level of cross-reactivity, future vaccines should attempt to further boost these antibodies, thus putatively enhancing protection against newly emerging influenza viruses.

## Material & Methods

### 2.1 Human serum samples

Human serum samples were obtained from participants enrolled in the Lifelines Biobank (Groningen, The Netherlands), a large, multigenerational, population-based cohort study comprising over 167,000 individuals from the northern Netherlands (18). The Lifelines protocol has been approved by the UMCG Medical Ethical Committee (2007/152). Blood samples were collected at two assessment timepoints (A1 and A2) from 180 healthy individuals, with an interval of approximately five years between collections (Fig. 1). Selected participants were stratified into three age cohorts at A1: adolescents aged between 17-18 (*n* = 60; 15 males and 45 females), adults aged between 37-41 (*n* = 60; 30 males and 30 females), and seniors aged between 62-67 (*n* = 60; 30 males and 30 females). For the adult and senior cohorts, A1 sampling occurred in 2009 and A2 in 2014. For the adolescent cohort, A1 occurred in 2011–2012 and A2 in 2015–2017 (see (17) for further details). Due to the absence of individual vaccination records, participants with chronic conditions such as asthma, cancer, or diabetes were excluded, as these individuals are more likely to have received additional influenza vaccinations in accordance with Dutch national guidelines. To assess the specificity and functional characteristics of nnABs in non-exposed individuals, we included from the originally 180 participants only those with a microneutralizing antibody (MNA) titer ≤40 against the A/California/7/2009 (H1N1pdm09) influenza virus at A1 (as determined in (4)). See Fig. 1 for a full overview of the included and excluded participant groups.

### 2.2 Preparation of Whole Inactivated Virus, HA subunit proteins, and nucleoprotein

The H1N1pdm09 influenza virus strain A/California/7/2009 was obtained from the National Institute of Biological Standards and Controls (NIBSC, Potters Bar, United Kingdom) and propagated in the allantoic fluid of embryonated chicken eggs, as previously described (31). Whole inactivated virus (WIV) was prepared by incubating the harvested and purified virus overnight with 0.1% (v/v) β-propiolactone (Acros Organics, Geel, Belgium) under continuous rotation at 4 °C. The inactivated virus was subsequently dialyzed overnight at 4 °C against HEPES-buffered saline (Thermo Fisher Scientific, Bleiswijk, The Netherlands) to remove residual β-propiolactone. Complete inactivation was confirmed by inoculating Madin-Darby Canine Kidney (MDCK) cells and assessing for viral replication using a hemagglutination assay (32). Subunit vaccine, primarily composed of hemagglutinin (HA), was derived from the corresponding WIV preparation following established protocols (33). Recombinant nucleoprotein (NP) was isolated from *Escherichia coli* strain AD494 after transformation with pET32a-NP plasmid (Merck Millipore) and extracted as described before (34). Total protein concentrations of WIV, purified surface proteins, and NP were quantified using the micro-Lowry assay (31).

### 2.3 Determination of influenza virus-specific IgG antibodies

Influenza-specific IgG responses targeting WIV, HA subunit (SU), and NP, as well as IgG subclasses (IgG1, IgG2, IgG3, and IgG4) specific to WIV were assessed using enzyme-linked immunosorbent assays (ELISAs), following previously described protocols (35). Neuraminidase (NA)-specific IgG titers had been determined previously by enzyme-linked lectin assay (17). ELISAs were executed as follows. Briefly, ELISA plates were coated with WIV (0.3 µg/well), HA subunit (0.1 µg/well), or NP (0.3 µg/well). Serum samples were pre-diluted 1:200 for WIV-, HA subunit-, and NP-specific IgG detection, and serially diluted up to 1:6400. On each plate, a serially diluted serum pool, obtained by combining sera from a collection of the Lifelines donors, was used as calibrator. The antibody levels of this serum pool were assigned a value of 100 arbitrary units (AU). If the OD value of a prediluted serum sample was lower than the OD value of the highest dilution of the calibrator the sample was assigned an AU value of 0. For the subtype-specific IgG ELISA, serum was prediluted 1:200 for IgG1 and IgG4, and 1:50 for IgG2 and IgG3. Horseradish peroxidase-linked goat anti-human IgG (SouthernBiotech, cat# 2040-05), mouse anti-human IgG1 (Thermofisher, A-10648), mouse anti-human IgG2 (Thermofisher, cat# MH1722), mouse anti-human IgG3 (Thermofisher, cat# MH1732), or mouse anti-human IgG4 antibodies (Thermofisher, cat# MH1742) were used to detect bound IgG, IgG1, IgG2, IgG3 and IgG4, respectively. Colorimetric development was carried out using O-phenylenediamine dihydrochloride (Sigma-Aldrich, USA), and absorbance was measured at 492 nm. AU values for individual samples were calculated using a logistic-log function for the readings of the serial diluted samples, as previously described (36).

### 2.4 FcγRIIIa-binding ELISA

A FcγRIII-binding ELISA was performed to assess the interaction between soluble FcγRIIIa and influenza-specific total IgG, as previously described (36). Briefly, ELISA plates were coated with WIV (0.3 µg/well), followed by incubation with serum samples pre-diluted 1:50 and applied in two-fold serial dilutions. All dilutions and washing steps were performed using PTG buffer (1× PBS supplemented with 0.02% v/v Tween-20 and 0.3% w/v gelatin; Merck, Germany). Biotinylated soluble FcγRIIIa (0.1 µg/mL) was added and incubated for 1 hour, followed by a 1-hour incubation with streptavidin-polymerized horseradish peroxidase (poly-HRP; Sanquin, cat# M2032) at 0.1 µg/mL. The assay was developed using O-phenylenediamine dihydrochloride (Sigma-Aldrich, USA), and absorbance was measured at 492 nm. FcγRIIIa binding was quantified in arbitrary units (AU), using a standard curve generated from serial dilutions of a pooled serum calibrator, as described for the IgG ELISA.

### 2.5 Natural killer cell isolation

NK cells were isolated from buffy coats obtained from Sanquin (Amsterdam, The Netherlands) using the RosetteSep™ Human NK Cell Enrichment Cocktail (Stemcell Technologies, Vancouver, Canada), following the manufacturer’s protocol. Briefly, 500 µl enrichment cocktail was added to 10 ml buffy coat and incubated at room temperature for 10 minutes. Next, the mixture was supplemented with 10 ml D-PBS and pipetted on 15 ml of Ficoll-Paque Plus (Cytiva, Wilmington, DE USA) in a 50 ml SepMate tube (Stemcell). After centrifuging the tube at 1200 x g at room temperature for 10 min, the supernatant containing the NK cells was collected. Before resting, the cells were washed twice with D-PBS 2% FBS. Isolated NK cells were resuspended in complete RPMI (cRPMI) medium (ThermoFisher Scientific) containing 10% heat-inactivated Fetal Bovine Serum (FBS; Life science production (LSP), Sandy, UK), 1% PSG (penicillin/streptomycin/L-glutamine 100x, ThermoFisher Scientific), and 5 ng/ml IL-15 (Biolegend, San Diego, CA, USA) at a final concentration of 1*10^6^ cells/ml (37). Cells were rested overnight at 37⁰C in a humidified incubator with 5% CO_2_.

### 2.6 Antibody-dependent cellular cytotoxicity assay

To evaluate the ability of influenza-specific antibodies to mediate ADCC, an antibody-dependent NK cell activation (ADNKA) assay was performed as previously described (37). ELISA plates were coated with WIV (0.3 µg/well) for 1 hour, blocked for 45 minutes with 2.5% milk powder (Protifar, Nutricia) diluted in 0.05 M carbonate-bicarbonate buffer (pH 9.6–9.8), and incubated for 1.5 hours with serum samples diluted 1:100. Wells without serum served as negative controls. After serum incubation, plates were washed five times with PBS and incubated overnight at 4 °C in PBS.

The following day, rested NK cells were washed once with RPMI and resuspended at 50,000 cells per well in 100 µL complete RPMI containing V450-labeled mouse anti-human CD107a antibody (1:50; BD Biosciences, Franklin Lakes, NJ, USA). Cells were incubated for 4 hours at 37 °C and 5% CO₂ in the serum-precoated plates. Post-incubation, cells were transferred to a 96-well round-bottom tissue culture-treated plate (Corning, NY, USA) and stained for 10 minutes on ice with ZombieNIR™ viability dye (1:1000; BioLegend) in FACS buffer (PBS + 2% v/v decomplemented FBS; Lonza). Subsequently, cells were stained with PE-labeled mouse anti-human CD56 antibody (1:50; BD Biosciences) for 30 minutes at 4 °C. After washing, samples were analyzed using a Quanteon™ flow cytometer (Agilent). Unstained cells were used to optimize voltage settings for each fluorophore, and Fluorescence Minus One (FMO) controls were employed to define gating strategies (supp. Fig. 5).

### 2.7 Statistics

Statistical analyses were performed using GraphPad Prism version 10 for Windows, GraphPad Software, Boston, Massachusetts USA, www.graphpad.com. The type of analysis used is indicated in the respective figure legend. Before performing the correlations, the distribution patterns of the various quantifications were assessed, and normalization was applied as needed. Specifically, FCγRIIIa-binding antibodies, total IgG (WIV), IgG subclasses, and antibody quantifications were normalized using log10-transformation. All correlation analyses and their corresponding visualizations were performed using the R programming language (38). Correlations were calculated using the *cor* function from the *base* stats package with the Pearson method. The results were visualized using the *corrplot* function from the *corrplot* package, and the corresponding significance levels associated with the correlations were determined using the *cor.mtest* function from the same package.

### 2.8 Ethics statement

The Lifelines study was approved by the ethics committee of the University Medical Center Groningen, document number METC UMCG METc 2007/152. Formal written informed consent was obtained from all individuals and parents of children included in the study.

## Data Availability

All data produced in the present study are available upon reasonable request to the authors

## Acknowledgements

The Lifelines Biobank initiative has been made possible by subsidy from the Dutch Ministry of Health, Welfare and Sport, the Dutch Ministry of Economic Affairs, the University Medical Center Groningen, University Groningen, and the Northern Provinces of the Netherlands. The authors wish to acknowledge the services of the Lifelines Cohort Study, the contributing research centres delivering data to Lifelines, and all the study participants. The authors would like to thank Federica Sicca, Eva Vermeulen, and Peter Laczo for excellent support in data collection.

